# Age-Related Characteristics of SYT1-Associated Neurodevelopmental Disorder

**DOI:** 10.1101/2025.05.14.25327512

**Authors:** Sam G. Norwitz, Josefine Eck, Joel S. Winston, Kate Baker

## Abstract

**Objectives:** We describe the clinical manifestations and developmental abilities of individuals with SYT1-associated neurodevelopmental disorder (Baker-Gordon syndrome) from infancy to adulthood. We further describe the neuroradiological and electrophysiological characteristics of the condition at different ages, and explore the associations between these characteristics and clinical symptoms.

**Methods:** Participants were recruited to the UK-based Brain and Behavior in Neurodevelopmental Disorders of Genetic Origin project. Caregivers completed a medical history questionnaire and a battery of standardized neurodevelopmental measures. MRI and EEG records were obtained with consent from treating clinicians. Age-related clinical manifestations and neuroimaging records were systematically analyzed. Accuracy testing was used to explore brain-symptom associations.

**Results:** This study describes 39 individuals with 29 distinct *de novo SYT1* missense variants, including 11 novel variants. Qualitative age-related clinical trends included the resolution of hypotonia and worsening of movement disorders, sleep difficulties, and self-injurious behaviors. Social-communicative impairments were prominent, with evidence of progression with age. MRI abnormalities were identified in 48% of individuals, while EEG abnormalities were present in 92%. Epileptiform activity frequently co-occurred with movement disorders, while irregular sleep EEG coincided with sleep difficulties and respiratory problems.

**Interpretation:** This study characterizes the broad spectrum and age-related progression of clinical symptoms and brain-related findings in individuals with SYT1-associated neurodevelopmental disorder. Further research is needed to understand factors contributing to within-individual change, and to develop targeted interventions aimed at improving outcomes and quality of life for affected individuals and their families.

## Introduction

Synaptotagmin 1 (SYT1) is an integral membrane protein that serves as a critical effector of presynaptic vesicle fusion, responsible for coupling activity-induced calcium (Ca^2+^) influx to fast, synchronous neurotransmission within the central nervous system ^1,2^. SYT1 is comprised of two cytoplasmic functional domains (C2A and C2B) containing negatively-charged aspartate residues that bind Ca^2+^ upon depolarization-induced influx at the nerve terminal. Ca^2+^ binding neutralizes these residues and induces a conformational change, which allows the protein to penetrate the negatively-charged plasma membrane, triggering vesicle fusion and release of neurotransmitters into the synaptic cleft ^2,3^. The *SYT1* gene is highly conserved across species, signaling its importance in neuronal function and neurodevelopment ^3,4^.

*De novo* missense variants in the *SYT1* gene result in functional impairment of presynaptic vesicle exocytosis ^5–7^. SYT1-associated neurodevelopmental disorder (SYT1-associated NDD; Baker-Gordon syndrome; OMIM #618218), arising from these variants, is characterized by infantile hypotonia, ophthalmic abnormalities, moderate to profound global developmental delay, hyperkinetic and involuntary movement disorders, and EEG abnormalities in the absence of overt seizures in most cases ^8^. With only 25 confirmed cases across the literature and no existing longitudinal or retrospective studies, there is limited information about the developmental trajectories and long-term prognoses of diagnosed individuals. Moreover, no studies to date have attempted to systematically describe the neuroradiological and electrophysiological characteristics of the disorder, and relate these characteristics to clinical evolution.

Since the first reported case of SYT1-associated NDD by our group a decade ago ^5^, the number of diagnosed individuals has increased considerably, and individuals previously reported have grown older. As such, we are now able to document symptom progression and neurobiological characteristics of SYT1-associated NDD, and examine their associations, with a view towards better understanding this rare disorder and strengthening post-diagnostic counselling and supportive care. In this paper, we retrospectively describe symptom manifestation and characterize the neuroradiological and electrophysiological features of the disorder across age bands from infancy through adulthood. We further explore the associations between neurobiological features and clinical symptoms.

## Methods

### Recruitment

Individuals with *SYT1* missense variants were recruited to the UK-based Brain and Behavior in Neurodevelopmental Disorders of Genetic Origin (BINGO) project. Study participants had received their genetic diagnosis within a local clinical service, following genome-wide sequencing (WES or WGS, trio or solo) by an accredited laboratory. All variants were confirmed *de novo*. Families were referred to BINGO following diagnosis by clinicians, or families self-referred via the project website. Written consent to share clinical and genetic data and collect phenotyping questionnaires for research purposes was obtained from parents or legal guardians under Cambridge Central Research Ethics Committee approval (IRAS 83633, REC ref: 11/0330/EE). Three additional individuals with *SYT1* missense variants were identified from published case reports ^9–11^, and openly-available information was included in the current paper.

### Data Collection

Lifetime medical history information was collected from parents/carers via a study-specific medical history questionnaire (MHQ) covering prenatal, infant, childhood and current health, as well as an inventory of developmental milestones. The MHQ was completed via an online form. For the three supplemental case reports, MHQ data was extracted from the published information where available. Parents/carers completed a battery of standardized neurodevelopmental questionnaires, with scoring and analysis procedures outlined in Supplementary Table 1. All questionnaires were completed online besides Vineland Adaptive Behavior Scales (VABS) assessments ^12^, which were completed via video interview (n=28) or telephone (n=1), based on parent preference. For parents/carers who did not have a working knowledge of English, a translator was provided by the research team and all questionnaires were completed via video interviews (n=6). Clinical MRI (n=55 reports from 29 individuals) and EEG (n=69 reports and 6 recordings from 30 individuals) records were provided by clinicians and/or hospitals with parent/carer consent.

### Data Analysis

Data were analyzed using MATLAB, version R2023b and R, version 4.4.2. Retrospective MHQ data was organized by age band (infancy [<1 year], childhood [1-10 years], and adolescence to adulthood [>10 years]). Where participants had partial datasets, all available data were included in analyses. Bar graphs were used to display symptom prevalence for each age band. Growth parameters were calculated in accordance with CDC growth chart percentile curves ^13^.

Raincloud plots were generated for each questionnaire measure. Where applicable, cut-off scores were overlaid to contextualize the severity of difficulties relative to normative populations or previously published data. To assess associations with age, questionnaire scores were analyzed using Spearman’s correlation. Results were visualized using a forest plot.

Kaplan-Meier survival analyses were used to model the ages at which participants achieved core developmental milestones. Analyses included participants who had either achieved the milestone or who were of an age when achieving each milestone could be expected, with lower bounds of inclusion based on the age at which 50% of typically developing children pass the milestone ^14,15^.

MRI reports were collated and grouped by age band as above. Reports were classified as either (1) normal or as reporting (2) developmental abnormalities and/or (3) acquired abnormalities, based on the classification schemes described by Ashwal et al. ^16^ and Robinson et al. ^17^. The latter two categories consisted of eight primary patterns of abnormality, including: (i) irregular skull morphology, (ii) delayed or immature myelination, (iii) cysts and masses, (iv) hypoplasia, (v) heterotopia, (vi) white matter lesions or abnormalities, (vii) cortical/subcortical atrophy, and (vii) hemorrhagic lesions. For individuals with multiple MRIs, serial longitudinal findings were compared to identify changes over time.

EEG records were collated and grouped by age band. All available reports and recordings were reviewed in consultation with a clinical neurophysiologist and classified according to the status of the following criteria: (i) overall abnormal, (ii) abnormal posterior background, (iii) epileptiform activity (wakeful and sleep-state), (iv) non-specific abnormalities (wakeful and sleep-state), (v) abnormal sleep features/architecture, (vi) superimposed abnormalities in sleep, (vii) abnormal response to photic stimulation, and (viii) events captured. Non-specific abnormalities were defined as irregular patterns of activity which were not characteristic of a particular etiology, and were classified as focal, multifocal, or generalized. Events were defined as clinically observed behaviors captured on EEG and were classified as epileptic or not. Reports and recordings with insufficient data or description to allow for confident overall categorization were excluded from the final analysis. For individuals with multiple EEGs, serial findings were compared to identify changes over time.

To explore brain-symptom associations, EEG/MRI findings were paired with age-relevant MHQ data. Accuracy-based classification testing was performed to cross-sectionally evaluate the predictive value of each neuroimaging feature for each concurrent clinical symptom. Accuracy (i.e., diagnostic effectiveness) was calculated as the proportion of true positive (co-occurrent neuroimaging features and clinical symptoms) and true negative (absent neuroimaging features and clinical symptoms) classifications relative to the total observations^18^. To account for repeated measures from participants with serial scans, a bootstrapping approach was employed, in which a random subset of scans was selected for accuracy testing, ensuring that only one observation per participant was included in each iteration. This process was repeated 500 times, and accuracy values were averaged across iterations to generate stable estimates that accounted for variability in participant selection while minimizing overfitting to a specific subset of data. The results were visualized using a heatmap displaying classification performance across feature pairs.

## Results

### Participants and Genetic Diagnoses

42 individuals with *SYT1* gene variants were recruited to the BINGO project. Inclusion criteria for the current analysis included a confirmed *de novo* missense *SYT1* variant without an alternative candidate variant more likely to explain the individual’s neurodevelopmental presentation. Based on these criteria, six individuals were excluded from the analysis (four with truncating variants, one with an inherited likely non-pathogenic variant, and one with an alternative pathogenic variant). An additional three study subjects who met the inclusion criteria were identified from published case reports ^9–11^. Thus, a total of 39 individuals were included in the final analysis. See Supplemental Table 2 for gene variant information.

### Physical and Behavioral Symptoms

At the time of assessment, participants included 2 (5.1%) infants, 25 (64.1%) children, and 12 (30.8%) adolescents and/or adults (Table 1). Retrospective data were available for all applicable prior age bands for every individual, except for one published case which lacked infancy data. Thus, data was available for a total of 38 individuals from infancy, 37 from childhood, and 12 from adolescence and/or adulthood.

**Table 1.** Participant demographics.

| **Demographics** | **N data available** | **Mean** | **Range** | **SD** |
| --- | --- | --- | --- | --- |
| **Age at time of participation, years** | 39 (100%) | 8.8 | 0.2 – 34.3 | 7.7 |
| Infant (< 1 year) | 2 (5.1%) | 0.5 | 0.16 – 0.92 | 0.5 |
| Child (1-10 years) | 25 (64.1%) | 5.0 | 1.2 – 8.4 | 2.2 |
| Adolescent (>10 years) | 12 (30.8%) | 17.9 | 10.8 – 34.3 | 7.4 |
| **Sex (female, no.)** | 19 (48.7%) |  |  |  |
| **Deceased (after assessment)** | 2 (5.1%) |  |  |  |
| **Pregnancy** | 35 (89.7%) |  |  |  |
| Pregnancy length, weeks | 33 (84.6%) | 39.1 | 34 – 42.5 | 2.1 |
| Pregnancy concerns | 18/33 (54.5%) |  |  |  |
| Reduced fetal movements | 8/33 (24.2%) |  |  |  |
| NICU/SCBU admission | 8/28 (28.6%) |  |  |  |
| **Growth parameters** |  |  |  |  |
| Birth weight, kg | 34 (87.2%) | 3.2 | 2.20 – 4.2 | 0.6 |
| Current weight, percentile | 22 (56.4%) | 23.8 | 0.4 – 97.8 | 29.2 |
| Current height, percentile | 21 (53.8%) | 33.2 | 0.9 – 99.8 | 31.9 |
| Orbitofrontal circumference, percentile | 22 (56.4%) | 32.7 | 0.6 – 99 | 31.2 |

#### Abnormal Muscle Tone

Hypotonia was highly prevalent in infancy (36/38 [94.7%]) and childhood (30/34 [88.2%]), but reduced in adolescence and adulthood (4/11 [36.4%]). A concomitant increase in hypertonia/stiffness was observed in six children and two adolescents, though the overall prevalence of hypertonia remained low (≤ 21%) across age bands.

#### Movement Disorders

The overall prevalence of movement disorders increased from 36% (8/22) in infancy to 85% (28/33) in childhood, and further increased to 100% (12/12) in adolescence and adulthood. Figure 1 shows a breakdown of movement disorder subtypes by age band. Hyperkinetic movement disorders were the most common across age bands, with little change in distribution from childhood onwards. Ataxia first emerged in childhood and persisted into adolescence/adulthood.

**Figure 1.**
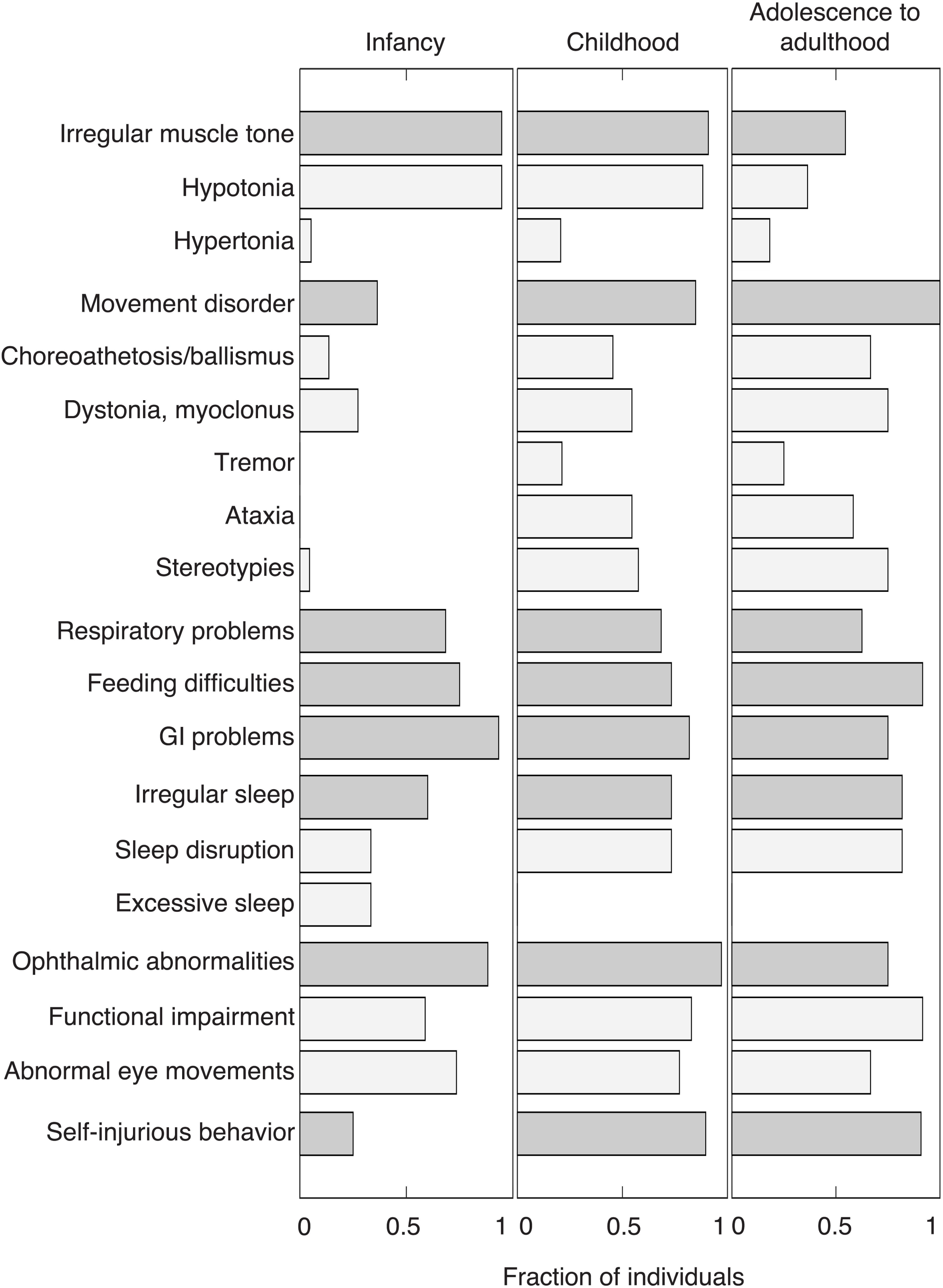
Clinical symptom prevalence by age band. Bar graphs depict the fraction of individuals across age bands who display each symptom. Age bands included infancy (<1 year), childhood (1-10 years), and adolescence to adulthood (>10 years). Dark grey bars represent overall prevalence for each symptom category; light grey bars represent subtype prevalence, if applicable.

#### Respiratory Problems

Respiratory problems were common from birth through adulthood without considerable change in overall prevalence. The most common respiratory problem was obstructive apnea, with aspiration, irregular breathing patterns and recurrent respiratory infections also common. Of the eight (29% [8/28]) newborns who required NICU/SCBU admission, seven were due to respiratory difficulties, with one mortality ^10^.

#### Feeding and Gastrointestinal (GI) Problems

Mechanical feeding problems, such as chewing, swallowing and sucking difficulties, were common without considerable change across age bands. Eight individuals required surgical procedures to assist with enteral feeding. Accompanying these feeding difficulties was a high prevalence of GI problems, which persisted across age bands. The most common GI problem in infants was gastroesophageal reflux disease, with irregular bowel motility also being common from childhood onwards.

#### Sleep Problems

In infancy, sleeping patterns varied widely, with 33% (10/30) reported to sleep excessively and 33% (10/30) having difficulties commencing and staying asleep. A substantial increase in sleep disruptions was observed in childhood (73% [22/30]), including the emergence of sleep disruptions in all the individuals who previously slept excessively, which persisted into adolescence and adulthood (82% [9/11]). Typical sleep disruptions were characterized by difficulty commencing sleep, frequent waking throughout the night, nighttime agitation, and extended periods without sleep.

#### Ophthalmic Abnormalities

Functional visual impairments and abnormal eye movements were highly prevalent across age bands. The most common functional impairment was cerebral visual impairment, present in 24% (8/34) of infants, 51% (18/35) of children, and 75% (9/12) of adolescents and adults. Issues with visual acuity, including hypermetropia and myopia, were also common, present in 29% (10/34) of infants, 54% (19/35) of children, and 50% (6/12) of adolescents and adults. The most common abnormal eye movement was strabismus, present in 50% (17/34) of infants, 57% (20/35) of children, and 50% (6/12) of adolescents and adults. Nystagmus was also common, present in 44% (15/24) of infants, 49% (17/35) of children, and 17% (2/12) of adolescents and adults.

#### Self-Injurious Behavior

Self-injurious behaviors—defined as repetitive, harmful actions directed towards one’s own body—were reported in only two infants but became highly prevalent in childhood (26/29 [90%]) and persisted into adolescence and adulthood (91% [10/11]). The most common form of self-injurious behavior was chewing one’s body parts (typically fingers and hands), followed by self-hitting, biting and head banging.

#### Developmental Milestones

The chronological ages at which individuals achieved developmental milestones are summarized in Figure 2. In all instances, the age of reaching developmental milestones was delayed when benchmarked to normative percentiles ^14,15^. Smiling emerged in all individuals, at a median age of 3 months. Sitting unsupported was attained by 80% (24/30; lower bound of inclusion = 8.2 months) at a median age of 18 months. Speaking in single words occurred in 48% (15/31; lower bound of inclusion = 10.1 months), with a median age of 2 years, while 30% (9/30; lower bound of inclusion = 20.6 months) progressed to speaking in sentences, with a median age of 4 years. Walking was achieved by 73% (22/30; lower bound of inclusion = 12.9 months) at a median age of 2.25 years. Daytime toilet training was achieved by 20% (5/25; lower bound of inclusion = 31.5 months for girls, 34.7 months for boys) at a median age of 3.5 years.

**Figure 2.**
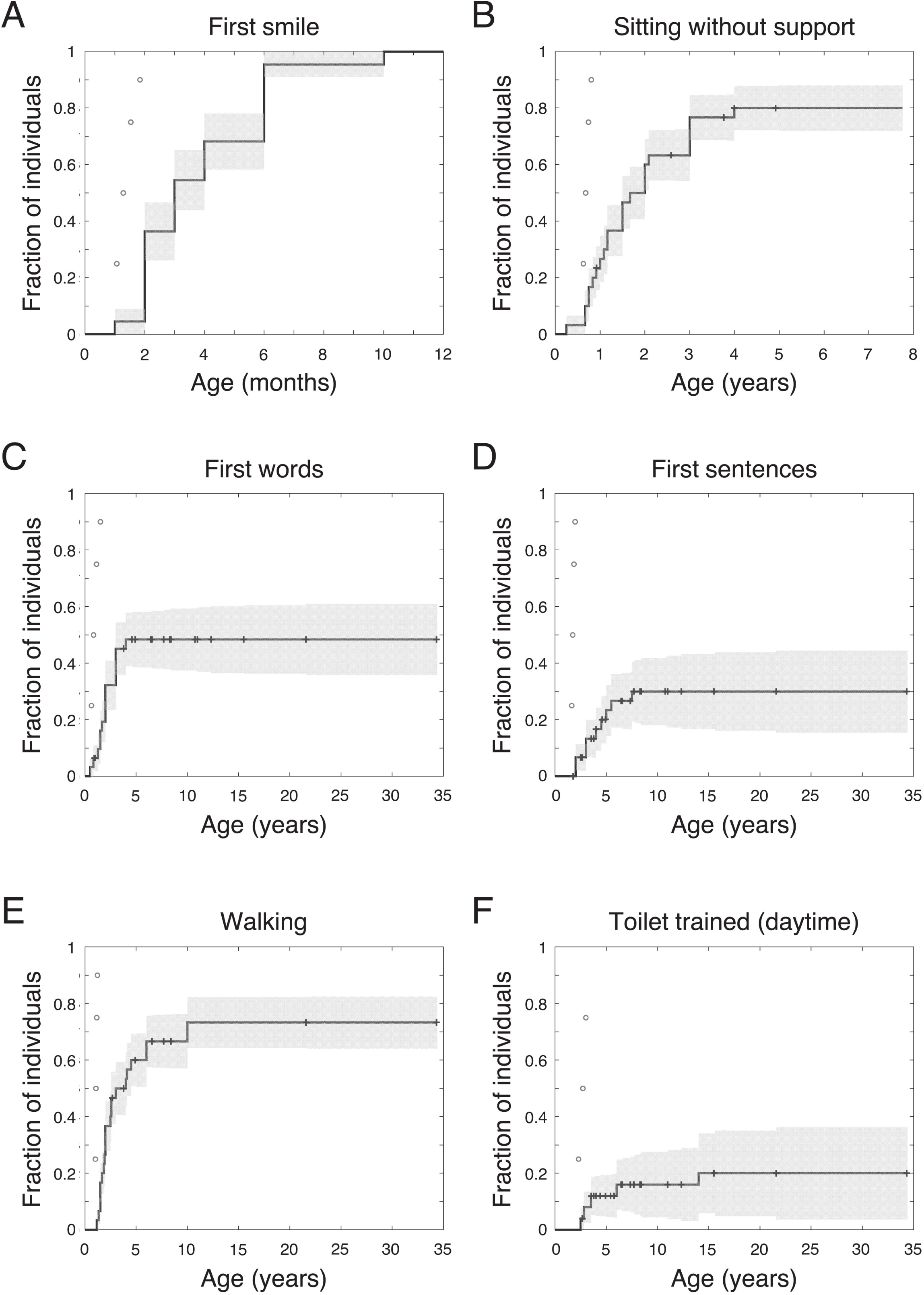
Acquisition of developmental milestones. Kaplan-Meier plots depict the fraction of participants who have achieved each milestone. Individuals who are of an appropriate age but who have yet to achieve each milestone are indicated by crosses with their age at the time of assessment. Normative percentiles for milestone acquisition are displayed as open circles.^14,15^

#### Neurodevelopmental Questionnaires

VABS Composite scores indicated a wide range of adaptive abilities, with 12% (3/25) of individuals falling within the normal/borderline range, 40% (10/25) classified as mildly to moderately impaired, and 48% (12/25) classified as severely to profoundly impaired. Among the VABS subdomains, communication was the most affected (Fig 3A).

**Figure 3.**
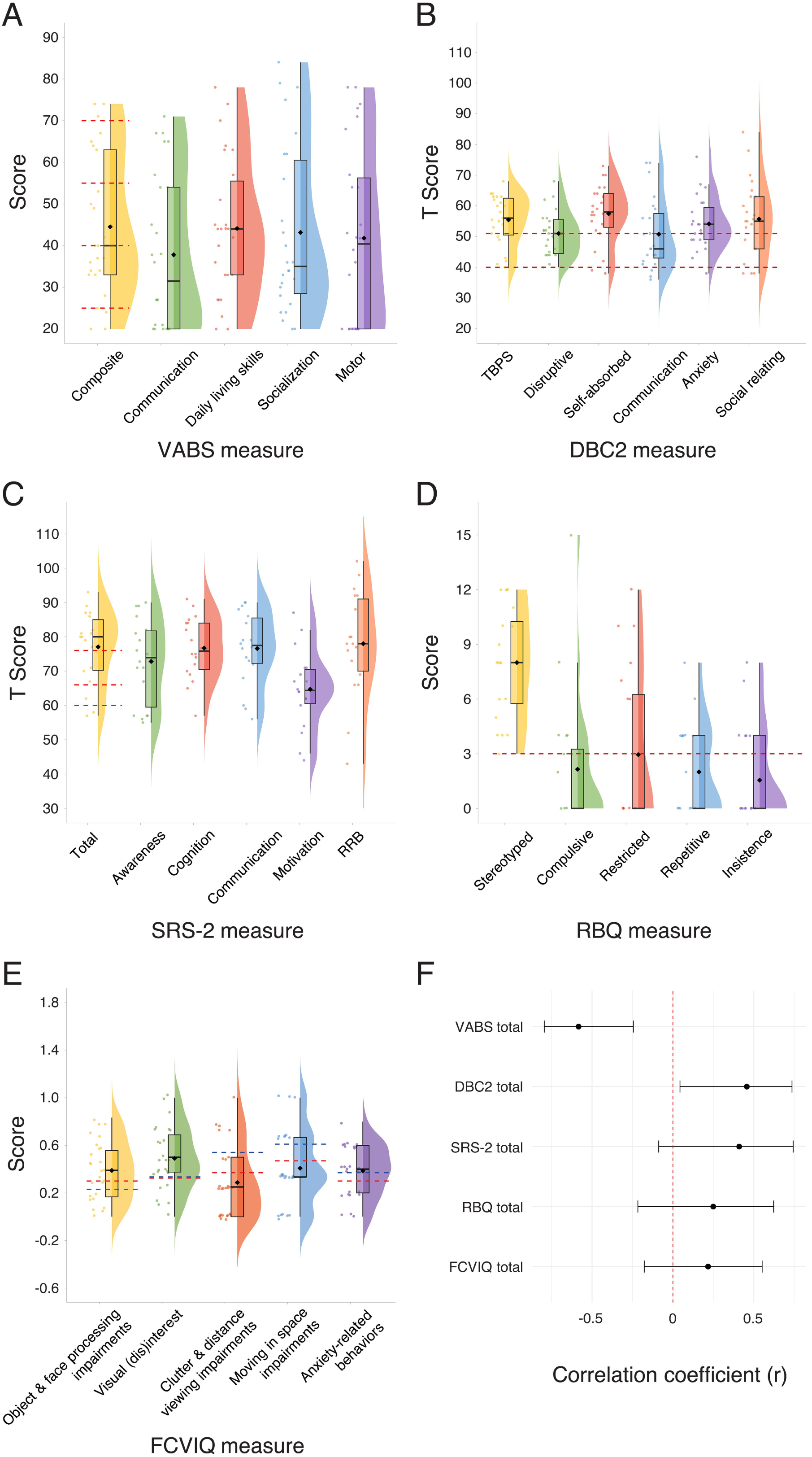
Neurodevelopmental Questionnaire Measure Scores. Violin plots display the distribution of scores on each of the questionnaire measures. Mean scores on each measure are represented by a black diamond. (A) Age-standardized Vineland Adaptive Behavior Scales (VABS). ID severity cut-offs are overlaid in red for the VABS composite score as follows: ‘likely unimpaired’ [top], ‘mild’, ‘moderate’, ‘severe’, ‘profound’ [bottom]. (B) Developmental Behavior Checklist (DBC2) T-scores. Levels of concern are overlaid in red for DBC2 subscales as follows: ‘serious’ [top], ‘moderate’, ‘little’ [bottom]. (C) Social Responsiveness Scale (SRS-2) T-scores. Severity cut-offs are overlaid in red for the SRS-2 total score as follows: ‘severe’ [top], ‘moderate’, ‘mild’, ‘normal’ [bottom]. (D) Restricted and repetitive Behavior Questionnaire (RBQ) Scores. Severity cut-offs are overlaid in red for all RBQ subscales as follows: ‘above’ [top], ‘normal’ [bottom]. (E) Flemish Cerebral Visual Impairment questionnaire (FCVIQ) factors scores. Mean scores for children with CVI are overlaid in red ^23^, and mean scores for children with CVI and unilateral are overlaid in blue^24^. (F) Forest plot displaying Spearman correlation coefficients (r) and 95% confidence intervals for the association between age and questionnaire measures. The dashed red line represents the null effect (r = 0).

The majority of Developmental Behavior Checklist (DBC2) ^19^ scores indexing emotional and behavioral difficulties were in the moderate to serious concerns range, with relatively greater difficulties with self-absorbed behavior—a DBC-2 subscale involving diverse items including mouthing objects, special interests and stereotyped behaviors (Fig 3B).

Social Responsiveness Scale (SRS-2) ^20^ scores indicated high levels of autism-associated traits, with severity varying within the atypical range. Social motivation emerged as a relative strength compared to other SRS-2 subscales (Fig 3C).

The Repetitive Behavior Questionnaire (RBQ) ^21^ assesses five domains of restrictive and repetitive behaviors. Most RBQ subdomains were within the normal threshold, except for stereotyped behaviors, which were markedly elevated (Fig 3D).

The Flemish Cerebral Visual Impairment Questionnaire (FCVIQ) ^22^ is a measure of behaviors associated with cerebral visual impairment (CVI) and its impact on everyday functioning. FCVIQ sum scores identified 81% (22/27) of individuals to be at risk for CVI (M = 4.70, SD = 1.27, factor sum range: 2-6). Factor scores on the FCVIQ revealed difficulties with object and face processing, visual interest, and anxiety-related behaviors of a level comparable to children with CVI ^23,24^ (Fig 3E).

Spearman correlation analysis revealed a significant negative association between participant age at time of data collection and VABS composite scores (rD = –0.58, n = 25, p = 0.002, 95% CI [–0.80, –0.24]), indicating lower global adaptive functioning in older individuals within the group. Additionally, a significant positive association was found between age and DBC scores (rD = 0.46, n = 22, p = 0.03, 95% CI [0.044, 0.74]), indicating more frequent or severe behavioral and emotional challenges in older individuals. Other measures also exhibited a trend toward greater difficulties with increasing age; however, these associations were not statistically significant (Fig 3F).

### Neuroradiology and Electrophysiology

A total of 55 MRI reports were obtained from 29 individuals. This included 24 scans from 20 infants, 22 scans from 18 children, and 9 scans from 5 adolescents and adults. 14 individuals had serial MRI scans across multiple age bands, enabling observation of within-participant change. The findings are summarized in Table 2. Sample MRI images are provided in Supplementary Figure 1.

**Table 2.** MRI classifications by age band.

| MRI Classification | Infants<br>(n=20<br>participants,<br>n=24 scans) | Children<br>(n=18<br>participants,<br>n=22 scans) | Adolescents &<br>Adults<br>(n=5<br>participants,<br>n=9 scans) | Total, MRI<br>feature ever<br>reported<br>(n=29<br>participants,<br>n=55 scans) |
| --- | --- | --- | --- | --- |
| Normal | 11 (55.0%) | 12 (66.7%) | 2 (40.0%) | 15 (51.7%) |
| Developmental abnormalities &<br>malformations | 8 (40.0%) | 6 (33.3%) | 3 (60.0%) | 13 (44.8%) |
| Irregular skull morphology | 6 | 4 | 1 | 8 |
| Delayed/immature myelination | 4 | 1 | 0 | 4 |
| Cysts and masses | 2 | 1 | 1 | 4 |
| Hypoplasia | 3 | 2 | 1 | 4 |
| Heterotopia | 0 | 1 | 1 | 2 |
| Acquired lesions | 2 (10.0%) | 4 (22.2%) | 2 (40.0%) | 7 (24.1%) |
| White matter lesions/ abnormalities | 1 | 3 | 1 | 4 |
| Cortical/subcortical atrophy | — | 3 | 2 | 5 |
| Hemorrhagic lesions | 1 | 0 | 0 | 1 |

Values indicate the number of individuals in each age band who display the feature. Values in the 'total' row indicate the total number of individuals who display the respective feature across age bands, accounting for individuals who are represented in multiple age bands.

The most prevalent developmental malformation was irregular skull morphology, most commonly microcephaly. Delayed myelination was common in infancy but typically resolved by childhood. Structural abnormalities included two left temporal arachnoid cysts, two non-specific focal masses, and hypoplasias of the septo-optic/pituitary region, hippocampal-fornix system, corpus callosum, insula, operculum, and posterior fossa. Acquired lesions consisted of persistent periventricular white matter lesions, and a choroid plexus hemorrhage in one infant which resolved in childhood. Progressive atrophy was observed for five individuals over serial scans and involved focal cerebellar and corpus callosal atrophy as well as three instances of diffuse cortico-subcortical atrophy with secondary ventricular dilation.

A total of 69 EEG reports and 6 recordings were obtained from 30 individuals. Upon review by a consultant neurophysiologist, 64 reports and 6 recordings from 26 patients had sufficient data for overall classification and between-subject comparison. This included 18 records from 13 infants, 38 records from 19 children, and 8 records from 6 adolescents and adults. The findings are summarized in Table 3. Sample EEG images are provided in Supplementary Figure 2.

**Table 3.** EEG classification by age band.

| EEG Classification | Infants<br>(n=13) | Children<br>(n=19) | Adolescents &<br>Adults<br>(n=6) | Total, EEG<br>feature ever-<br>reported<br>(n=26) |
| --- | --- | --- | --- | --- |
| Wakeful scans captured | 12 (92.3%) | 19 (100%) | 6 (100%) | 26 (100%) |
| Overall abnormal | 11/12 | 18/19 | 5/6 | 24/26 |
| Abnormal posterior background | 7/8 | 11/13 | 3/5 | 16/21 |
| Epileptiform activity | 5/9 | 13/18 | 4/6 | 18/26 |
| Non-specific abnormalities | 10/10 | 15/16 | 4/5 | 21/23 |
| Sleep scans captured | 4 (30.8%) | 9 (47.4%) | 2 (33.3%) | 13 (50.0%) |
| Abnormal sleep features | 1/3 | 2/7 | 0/2 | 3/10 |
| Superimposed abnormalities | 4/4 | 9/9 | 1/2 | 12/13 |
| Epileptiform activity | 2/4 | 8/9 | 1/2 | 9/13 |
| Non-specific abnormalities | 4/4 | 5/7 | 1/2 | 8/10 |
| Abnormal photic stimulation | 1/4 (25.0%) | 1/5 (20.0%) | 0/3 (0%) | 2/11 (18.2%) |
| Events captured | 3* | 3 | 0 | 4 |

Wakeful states were captured across 62 studies for 26 individuals. The archetypal wakeful pattern consisted of abnormal posterior background rhythms, primarily characterized by bursts of synchronous high-voltage polymorphic slow waves. Focal and/or multifocal epileptiform activity was commonly observed (22/26 [86%]), with a tendency towards maximal activity in bilateral posterior regions. Importantly, such epileptiform activity was evident without any associated behavioral manifestations suggesting a clinical seizure. Sleep was captured in 27 recordings across 13 individuals. Physiological sleep architecture was largely normal. Epileptiform activity, characterized by high amplitude spike-wave complexes and sharp waves, enhanced in sleep and tended to increase and spread with slow wave sleep. Of the 4 individuals without observed epileptiform abnormalities, 3 displayed non-specific abnormalities during sleep, with both focal and generalized localization. Where commented upon, abnormal paroxysmal patterns on sleep EEG were not associated with observations suggestive of clinical seizure.

11 individuals had serial EEGs across multiple age bands. Of these, EEGs remained overall abnormal for 10 individuals and became abnormal for one. Wakeful epileptiform activity emerged in three individuals and persisted in six. Two individuals had serial sleep studies, both of which remained abnormal with epileptiform activity and superimposed abnormalities.

### Clinical and Neurobiological Correlates

Accuracy testing on all available cross-sectional data, including bootstrapping to account for multiple observations of individuals across age bands, (Fig 4) revealed that abnormal MRI findings were concurrent with clinical features with 51% accuracy on average, while abnormal wakeful EEG and sleep EEG findings had average accuracies of 72% and 66%, respectively. That is, neuroimaging findings were concurrent with clinical symptoms in that percentage of instances, considering correct identification of symptom presence or absence. More specifically, abnormal MRI findings were concurrent with irregular sleep with 71% accuracy, including 69% accuracy specifically for developmental malformations. Among abnormal wakeful EEG features, epileptiform activity co-existed with movement disorders with the highest accuracy (93%). Abnormal sleep EEG findings were concurrent with irregular sleep patterns with 100% accuracy and respiratory problems with 94% accuracy, with epileptiform activity in sleep predicting both with 93% accuracy.

**Figure 4.**
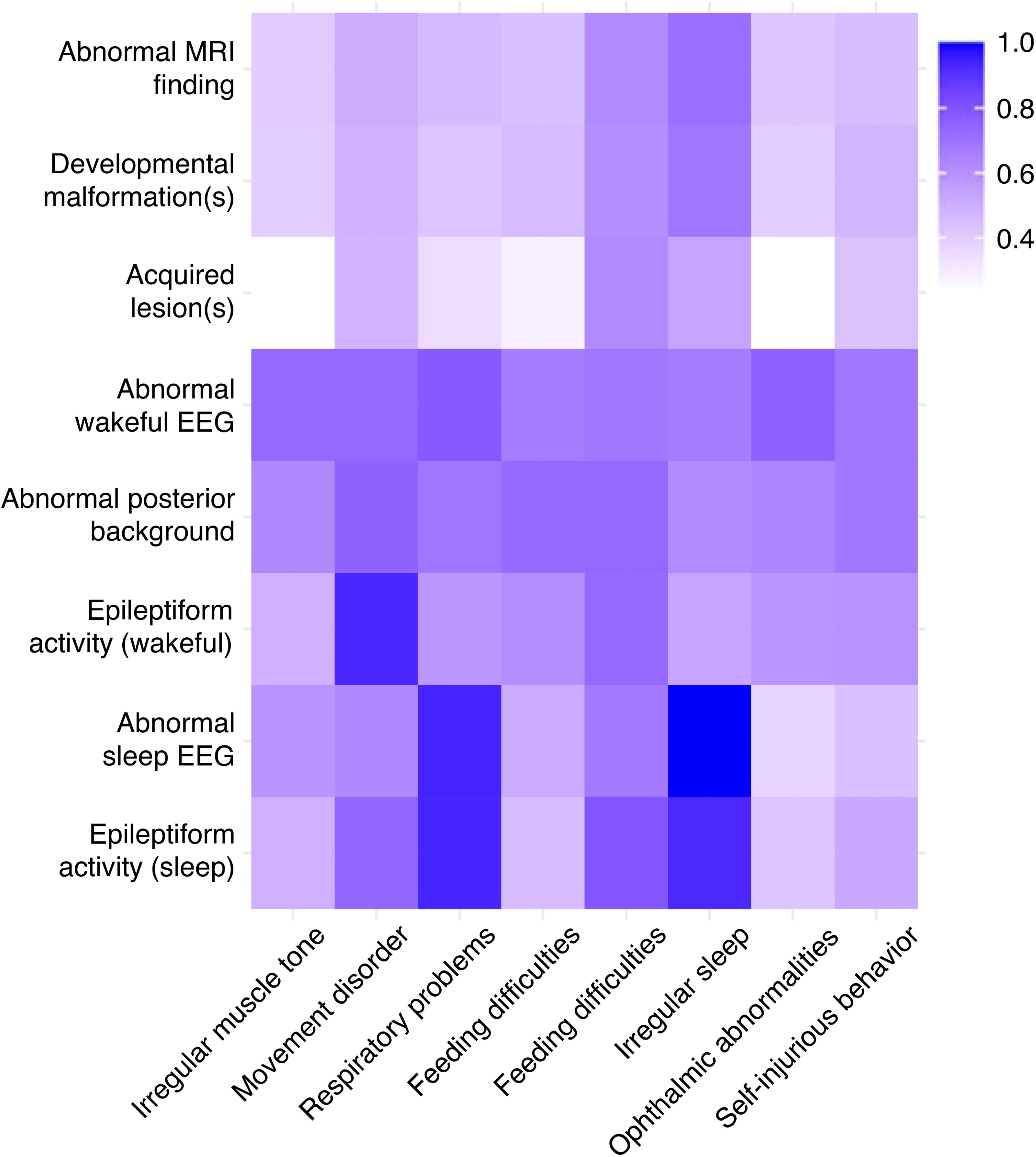
Accuracy heatmap for neuroimaging features clinical symptoms. Heatmap depicting classification accuracy for EEG/MRI features paired with concurrent clinical symptoms. Darker shades indicate higher classification accuracy.

## Discussion

In this study, we retrospectively mapped age-related physical and behavioral symptoms in 39 individuals with SYT1-associated NDD. We further analyzed 125 MRI and EEG records across 30 individuals, assessing age-related characteristics and relation to clinical presentation. The clinical features associated with *SYT1* missense variants in the current study are consistent with previous smaller case series ^8^; however, age-related trends and associations with neurobiological characteristics can now be observed for the first time.

Age-band analysis indicated that hypotonia often resolves with age, and, in some cases, is associated with a concomitant increase in hypertonia and spasticity. Similar dynamic shifts in muscle tone have been observed in infantile hypotonia of heterogeneous origins, with contributions from genetic disorders affecting key structural proteins of the nervous system^25^. Since muscle tone is essential not only for gross motor development but also for smooth muscle function, hypotonia may contribute to respiratory distress and feeding difficulties in infants and children with *SYT1* variants ^25,26^.

Another age-related trend was the increasing prevalence of involuntary movement disorders, primarily of the hyperkinetic type. Given that SYT1 is the primary isoform facilitating terminal dopamine release in the midbrain, its dysfunction likely contributes to the predominance of basal ganglia-related movement disorders, including chorea-athetosis, ballismus, dystonia, myoclonus, and stereotypies ^2,27^. The progression of motor symptoms may reflect a combination of mechanisms, including progressive synaptic dysfunction, maladaptive plasticity and reorganization, and the failure of compensatory mechanisms within the cortico-basal ganglia-thalamic circuitry ^28^. However, we also observed considerable variation in the types and severity of movement abnormalities within the group, for reasons that are not yet understood. Future studies should explore whether SYT1 variants lead to progressive synaptic dysfunction, and potentially age-related neuronal loss, disrupting the fine-tuning of motor pathways and exacerbation of hyperkinetic movement disorders with age.

Retrospective assessment of sleep habits revealed a developmental pattern characterized by excessive sleepiness in infancy followed by the emergence of severe sleep disturbances in childhood, with frequent waking and extended periods without sleep (sometimes lasting days). Sleep disorders are highly prevalent in individuals with pervasive NDDs, affecting up to 86% of cases ^29,30^. Potential contributors include hormonal imbalances, neurotransmitter dysregulation, medical comorbidities (e.g., respiratory issues, GI dysfunction, epilepsy), medication effects, sensory sensitivities and environmental stressors ^29^. Sleep disturbances in NDDs are also strongly linked to behavioral and psychological factors, including lower adaptive function, increased internalizing and externalizing behaviors and greater autism symptom severity ^31^. The relationship is bidirectional, as children with NDDs may be more vulnerable to sleep disturbances due to underlying biological and behavioral factors, while chronic sleep deprivation exacerbates neurodevelopmental challenges ^31–34^. In individuals with SYT1-associated NDD, the evolution of sleep problems may reflect a convergence of changes in medical comorbidities (e.g., respiratory and gastrointestinal dysfunction, epileptiform activity), medication use, and the progression of cognitive and behavioral differences, all of which likely contribute to the observed developmental shift from excessive sleepiness in infancy to disrupted sleep patterns in childhood and adolescence.

In line with our previous research, we found considerable variability in the acquisition of developmental milestones and adaptive functioning, with most individuals showing mild to profound impairments, particularly in communication ^8^. Building on these findings, standardized questionnaire measures highlight variably severe social-emotional-behavioral concerns including pronounced self-absorbed behaviors, anxiety, and autism-related traits. Stereotyped behaviors, self-injurious behaviors, and poor functional vision (CVI) were also reported to be prevalent within the SYT1 group, highlighting the challenge of discriminating between motor or sensory problems versus cognitive or behavioral characteristics. Cross-sectional analysis highlighted the potential for worsening adaptive functions with age, however this should be interpreted with caution because of the differences in ascertainment for genome-wide testing over time (i.e. younger-diagnosed individuals may be from a less severely impacted population on average) and previously observed decline in scores on age-standardized measures within NDD populations ^35^.

While previous literature on SYT1-associated NDD has reported predominantly normal MRI findings ^6,8–10^, the current study found that half of the individuals exhibited some structural abnormalities. Many of these were incidental or of limited functional significance, such as irregular skull morphology or delayed myelination, which largely stabilized or resolved with age. However, in some individuals, serial scans revealed evidence of progressive cortico-subcortical atrophy. Additional follow-up imaging will be crucial to determine whether these progressive changes are a common feature of the disorder, and whether this reflects clinically-relevant pathology.

In line with prior reports ^6,8,9^, EEGs collated for this *SYT1-*related NDD cohort consistently displayed widespread abnormalities. These were characterized by delayed background activity interspersed with bursts of synchronous high-voltage slow waves, predominantly in posterior regions, alongside isolated epileptiform discharges. In this study, we found largely preserved sleep architecture but a high prevalence of epileptiform activity, exacerbated by slow-wave sleep. Analysis of EEG features over time highlighted a general increase in abnormalities, whilst cross-correlating EEG and clinical characteristics highlighted co-occurrence between EEG abnormalities and movement disorders, respiratory problems and sleep disruptions with high accuracies. Epileptiform activity and movement disorders frequently co-occur in genetic neurodevelopmental conditions due to shared genetic mechanisms, with presentation patterns and temporal relationships varying by underlying genetic etiology ^36–38^. Likewise, the relationship between sleep disruption and epileptiform activity emerging during childhood and adolescence is well-documented across multiple genetic syndromes ^39,40^. An established dynamic interplay exists between these linked features and respiratory dysfunction. Sleep-disordered breathing—particularly obstructive apnea, the most common respiratory symptom observed in this study—exacerbates discharge frequency and severity, and conversely, can be influenced by epileptic activity ^41,42^.

Notably, in SYT1-associated NDD cases, overt behavioral manifestations or clinical seizures have very rarely been observed ^6,8^, despite the presence of epileptiform discharges during both wakefulness and sleep. This differentiates the condition from most other synaptic vesicle cycling disorders ^43^ and SNAREopathies ^44^. The neuronal origins, functional significance and clinical implications of these EEG observations remain an important area for future research, as subclinical epileptiform activity has been linked to cognitive impairment and decline across various neurological and psychiatric disorders ^45,46^. It is plausible that the heightened level of cortical hyperexcitability and hypersynchronization of neuronal networks associated with EEG oscillations and epileptic activity could underlie impaired gating and modulation of motor commands, regulation of autonomic functions such as respiratory rate and sleep cycles, and neural systems functions relevant to cognition. Together, these findings underscore the need for deeper mechanistic understandings of the multisystem interactions in SYT1-associated NDD to inform more precise clinical strategies for managing symptoms and promoting adaptive outcomes.

The current study has several limitations. First, given that each participant was diagnosed and recruited at a different age, some individuals were missing information from prior age bands. Second, data for the three individuals who were identified through published case reports were not standardized. Third, multiple clinicians contributed to MRI and EEG reporting, which may have introduced variability and dataset inconsistencies. Fourth, while accuracy testing provided a preliminary exploration of brain–symptom co-occurrence in our small sample size, its descriptive nature precludes causal interpretation, as associations may be confounded by other factors and results may be biased by outcome imbalances (i.e., when certain features are simply more common or rare, or demonstrate similar age-related trajectories). Lastly, SYT1-associated NDD is a rare syndrome with approximately only 50 confirmed cases worldwide. It is possible that milder or less typical forms of the disorder remain undiagnosed, as ascertainment for genomic testing varies globally and continues to change over time. As more clinicians become aware of the syndrome, and whole genome sequencing becomes more widely available, more cases will be identified, and different clinical patterns may become apparent. The current case series expands the catalogue of *de novo* missense *SYT1* variants, including 11 variants not previously reported (Supplementary Table 2). Functional analysis is not yet available for all variants, hence pathogenicity remains uncertain for some. The effects of variant type, location and functional impact on clinical evolution over time were beyond the scope of this study and should be a future priority topic.

In conclusion, the current study presents the most comprehensive overview to date of the clinical, behavioral, neuroradiological, and electrophysiological features of SYT1-associated NDD. We characterized the broad spectrum of clinical symptomology and neuroimaging features in individuals with the disorder, including how they change over time from infancy through adulthood. Further research is needed to deepen our understanding of these developmental changes and to develop targeted interventions aimed at improving outcomes and quality of life for affected individuals and their families.

## Acknowledgements

We appreciate the important contributions of each individual with an SYT1 variant and their families and caregivers. We also acknowledge the essential roles of the clinicians and laboratory scientists involved in the diagnostic journey and ongoing support of each case. This work was supported by the Medical Research Council (MC_UU_00030/3), Great Ormond Street Hospital Charity, Cambridge NIHR Biomedical Research Centre, and Gates Cambridge.

## Author Contributions

S.G.N. and K.B. contributed to conception and design of the study. S.G.N. and J.E. contributed to the acquisition of clinical data; J.S.W. and S.G.N. contributed to the review of neuroimaging/EEG records. S.G.N. contributed to data analysis. S.G.N. and K.B. contributed to drafting the text and figures, and all authors revised and approved the submitted manuscript.

## Data Availability

Reporting of SYT1 variants in open access repositories is listed in Supplemental Table 2. Anonymized data may be made available to other researchers from the corresponding author on reasonable request, and on condition of signing a Code of Conduct guaranteeing that the data will be kept confidential and securely.

