## Supplemental materials for "Age-Related Characteristics of SYT1-Associated Neurodevelopmental Disorder"

### Supplementary Materials for: Age-Related Characteristics of SYT1-Associated Neurodevelopmental Disorder

[**Supplementary Table 1.** Neurodevelopmental questionnaire measures](#_Toc177641311)	2

#### **Supplementary Table 1.** Neurodevelopmental questionnaire measures.

| Questionnaire | Description | Scoring Procedure |
| --- | --- | --- |
| Medical History Questionnaire (MHQ) | The MHQ assesses the following symptoms from infancy, childhood, and the past six months: abnormal muscle tone, movement disorders, feeding difficulties, sleep difficulties, sensory impairments, and self-injurious behavior. Other measures include prescriptive medication, psychiatric diagnoses, and educational/occupational placement. | Symptoms were classified as either being present or absent to determine overall prevalence. Adolescent and adult data was taken from the ‘past six months’ category, if the individual was ≥ 10 years 6 months at the time of assessment. |
| The Vineland Adaptive Behavior Scales (VABS) (Third Edition, Parent/Caregiver Form, Vineland-3) ^1^ | The VABS is a standardized assessment of global adaptive ability, comprised of three subdomains (communication, daily living skills, and socialization) and a separate measure of motor skills. | Where multiple assessments were available, most recent assessments were used. Raw VABS composite and subdomain scores were scaled according to published norms. For each domain, results are expressed as age standardized scores with a mean of 100 and standard deviation of 15 (i.e., a normal score is between 85 and 115). The VABS composite was categorized as ‘normal/borderline’, ‘mild/moderate’, or ‘severe/profound’, and was used here as a proxy measure of severity of developmental delay or intellectual disability (DD/ID). |
| Developmental Behavior Checklist (DBC2) ^2^ | The DBC2 is a standardized measure of emotional-behavioral difficulties in individuals with ID comprised of 5 subscales (disruptive/antisocial behavior, self-absorbed behavior, social relating, communication disturbance, and anxiety). | T-scores for the total behavioral problems score (TBPS) and all five treatment subscales were generated using the Australian norms. T-scores were not stratified for severity of ID due to missing VABS composite scores. |
| Social Responsiveness Scale (SRS-2) ^3^ | The SRS-2 is a screening tool designed to assess autism characteristics in the general population, and is comprised of 5 subscales (social awareness, social cognition, social communication, social motivation, and restricted interests and repetitive behavior). | SRS-2 raw scores were converted to age- and sex-appropriate T-scores based on published norms. |
| Repetitive Behavior Questionnaire (RBQ) ^4^ | The RBQ is a measure of repetitive behavior frequency across 5 subscales (stereotyped behavior, compulsive behavior, insistence on sameness, restricted preferences, and repetitive use of language). | Scores on the RBQ were calculated using the verbal scoring approach and missing items were scored as 1 (never). |
| Flemish Cerebral Visual Impairment Questionnaire (FCVIQ) ^5^ | The FCVIQ is a measure of behaviors associated with CVI and its impact on everyday functioning. It gauges 6 domains and 9 subscales of functional visual impairment (visual attitude [fixation, visual field, visual attention, influence familiar environment], ventral stream, dorsal stream, complex visuomotor abilities, other senses, and associated characteristics). | The analysis here focused on total scores. For descriptive purposes, we generated the Sum Score described by Ortibus et al.^5^ Using this score, an individual is considered at risk of CVI if they display a minimum of one behavior in at least four of the FCVIQ domains. We also generated scores for the five factors identified by Ben Itzhak et al. ^6^ (object and face processing impairments, visual (dis)interest, clutter and distance viewing impairments, moving in space impairments, and anxiety-related behaviors), which we scored according to the method described by Crotti et al. ^7^. Higher scores on all FCVIQ measures reflect greater impairment. |

#### **Supplementary Table 2.** SYT1 gene variant information. Nucleotide and amino acid changes for variants included in the current study are listed along with their classification, as listed on patients’ genetic reports and on prior publications ^8,9^. Bolded rows represent new variants not previously reported on. All variants are in relation to reference sequence: NM_005639.3.

| **Nucleotide and Amino Acid Change** | | **Domain & Type** | **Pathogenicity on report** | **Pathogenicity based on GiM and eBioMed papers** | **Pathogenicity as currently listed in ClinVar** | **n** |
| --- | --- | --- | --- | --- | --- | --- |
| c.476T>G | p.Leu159Arg(L159R) | C2A missense | Possibly pathogenic | Uncertain significance (GiM); pathogenic (eBioMed) | Likely pathogenic | 1 |
| c.551 T>C | p.Val184Ala(V184A) | C2A missense | Likely pathogenic | - | Likely pathogenic | 1 |
| c.587C>A | p.Thr196Lys(T196K) | C2A missense | Possibly pathogenic | Uncertain significance (GiM); pathogenic (eBioMed) | - | 1 |
| c.625G>A | p.Glu209Lys(E209K) | C2A missense | Possibly pathogenic | Uncertain significance (GiM); pathogenic (eBioMed) | - | 1 |
| c.655G>C | p.Glu219Gln (E219Q) | C2A missense | Possibly pathogenic | Uncertain significance (GiM); pathogenic (eBioMed) | Uncertain significance | 1 |
| c.907A>G | p.Met303Val(M303V) | C2B missense | Likely pathogenic | Likely pathogenic (GiM); pathogenic (eBioMed) | - | 1 |
| c.908T>A | p.Met303Lys (M303K) | C2B missense | Likely pathogenic | Pathogenic (GiM) | Pathogenic | 1 |
| **c.910G>C** | **p.Asp304His(D304H)** | C2B missense | Uncertain significance | - | - | 1 |
| c.911A>G | p.Asp304Gly(D304G) | C2B missense | Possibly pathogenic | Pathogenic (GiM) | Pathogenic | 1 |
| **c.920G>A** | **p.Gly304Asp(G307D)** | C2B missense | Pathogenic | - | Likely pathogenic | 1 |
| c.925T>C | p.Ser309Pro(S309P) | C2B missense | Likely pathogenic | Pathogenic (GiM / eBioMed) | - | 1 |
| **c.926C>T** | **p.Ser309Phe(S309F)** | C2B missense | Probably pathogenic | - | Pathogenic /  Likely pathogenic | 1 |
| **c.928G>A** | **p.Asp310Asn(D310N)** | C2B missense | Unclear significance | - | Likely pathogenic | 1 |
| **c.935A>G** | **p.Tyr312Cys(Y312C)** | C2B missense | Unknown significance | - | Likely pathogenic | 1 |
| **c.989_991delCAA** | **p.Thr330del(T330del)** | C2B deletion | Probably pathogenic | - | - | 1 |
| NA | **p.Asn341Lys(N341K)** | C2B missense (homozygous) | Possibly pathogenic | - | - | 1 |
| c.1022A>G | p.Asn341Ser (N341S) | C2B missense | Probably disease-causing | Uncertain significance (GiM) | Uncertain significance | 1 |
| c.1094A>G | p.Tyr365Cys(Y365C) | C2B missense | Likely pathogenic | Likely pathogenic (GiM); pathogenic (eBioMed) | - | 1 |
| **c.1095_1097dup** | **p.Asp366dup(D366dup)** | C2B duplication | Likely pathogenic | - | - | 1 |
| **c.1096G>A** | **p.Asp366Asn(D366N)** | C2B missense | Likely pathogenic | - | - | 1 |
| c.1098C>A  c.1098C>G | p.Asp366Glu (D366E) | C2B missense | Possibly pathogenic | Pathogenic (GiM) | Pathogenic | 2  1 |
| c.1100_1102dup | p.Lys367dup(K367dup) | C2B duplication | Likely pathogenic | Likely pathogenic (GiM) | Likely pathogenic | 1 |
| c.1101_1103dup | p.Lys367_Ile368insMet  (K367_I368insM) | C2B duplication, insertion | Pathogenic | - | Likely pathogenic | 1 |
| NA | **p.Ile368Ser(I368S)** | C2B missense | Possibly pathogenic | - | - | 1 |
| c.1103T>C | p.Ile368Thr(I368T) | C2B missense | Pathogenic | Pathogenic (GiM) | Pathogenic | 8 |
| c.1106G>A | p.Gly369Asp(G369D) | C2B missense | Possibly pathogenic | Likely pathogenic (GiM); pathogenic (eBioMed) | - | 1 |
| c.1113C>G | p.Asn371Lys(N371K) | C2B missense | Possibly pathogenic | Pathogenic (GiM) | Pathogenic | 2 |
| **c.1199G>A** | **p.Arg400Gln(R400Q)** | C2B missense | Uncertain significance | - | - | 1 |
| c.1202C > T | p.Pro401Leu(P401L) | C2B missense | Likely pathogenic | - | - | 1 |
| ***11 novel variants not previously reported on** | | **5 C2A**  **34 C2B** |  |  |  | **39** |

#### **Supplementary Figure 1.** Sample MRI images.

#### (A) Unremarkable axial and sagittal MRI.

#### (B) Axial T1-weighted and FLAIR MRI displaying atypical periventricular white matter lesions and suspected heterotopia.

#### (C) Serial axial T2 FLAIR MRIs from a patient during infancy (left) and early childhood (right) showing deep white matter asymmetry associated with left frontal corico-subcortical atrophy along with secondary dilation of the supratentorial ventricular system.

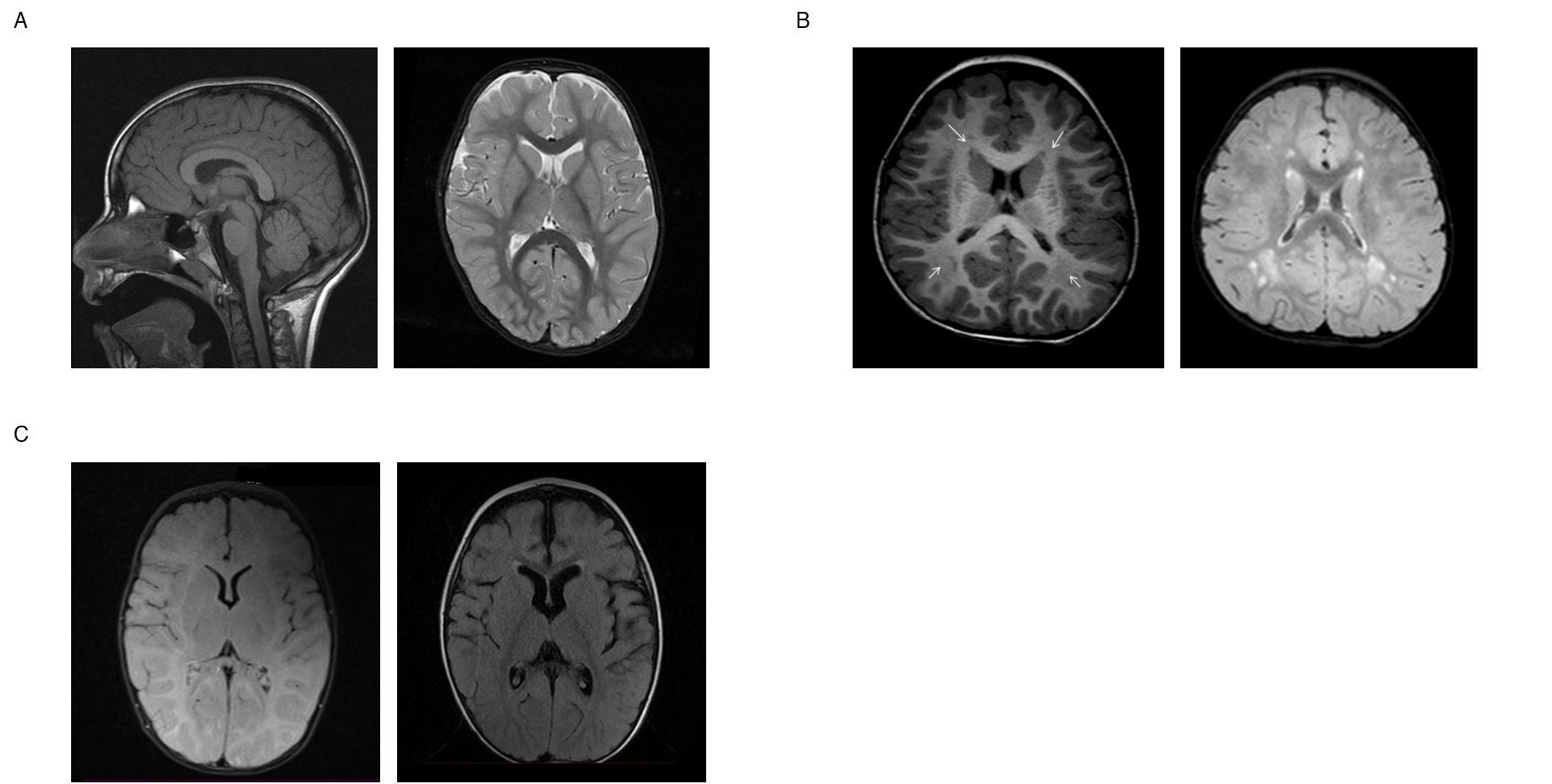

#### **Supplementary Figure 2.** Sample EEG images.

#### (A) Serial EEGs from a patient during early childhood, mid-childhood and early adolescence, showing an evolution in runs of widespread rhythmic theta/delta activities with posterior temporal rhythmic spiking (particularly in the earlier record) and variable associated epileptiform complexes. Left and right subpanels are shown on a common average referential montage; center subpanel on an AP bipolar montage with left-right alternation. Note that the timebase varies across the panels.

#### (B) Serial EEGs from a patient during infancy, early childhood, and adolescence displaying high amplitude theta-delta activity with frequent sharp and slow wave complexes. Left and center panel shown on an AP bipolar montage; right subpanel shown on a common average referential montage. Note that the timebase varies across the panels.

#### (C) EEG from a patient during early childhood displaying rhythmic slow and sharp waves often in runs of 8-14 sec, with a parieto-temporal maximum (right more than left). Shown on an AP bipolar montage.

#### (D) EEG from a patient during early childhood displaying high amplitude rhythmic theta/delta (3-4/s) activity predominately in the posterior regions. Shown on an AP bipolar montage.

#### (E) EEG a patient during early childhood displaying a flexor spasm with some electrodecrement. Interictal EEG shows frequent sharp waves/spikes. Shown on an AP bipolar montage.

#### (F) EEG from a patient during early childhood displaying a pattern interpreted as a subclinical seizure including rhythmic spike-wave complexes with posterior maximum. Shown on an AP bipolar montage.

#### (G) EEG from a patient during early childhood displaying some rhythmic delta activity with a posterior maximum and generalized slowing without clear epileptiform activities. Shown on an AP bipolar montage.

#### (H) Sleep EEG from an infant patient displaying synchronous high-voltage slow waves and traces of spindles. Shown on an AP bipolar montage.

#### (I) Serial sleep EEGs from a patient during early childhood, mid-childhood and early adolescence displaying brief runs of high amplitude fast spike and slow wave complex discharges over fronto-central regions and intermittent generalized bursts of high amplitude theta (5/s) activities. Shown on a common average referential montage (left and right subpanels) or an AP-bipolar montage (center panel). Note subtle differences in timebase between panels.

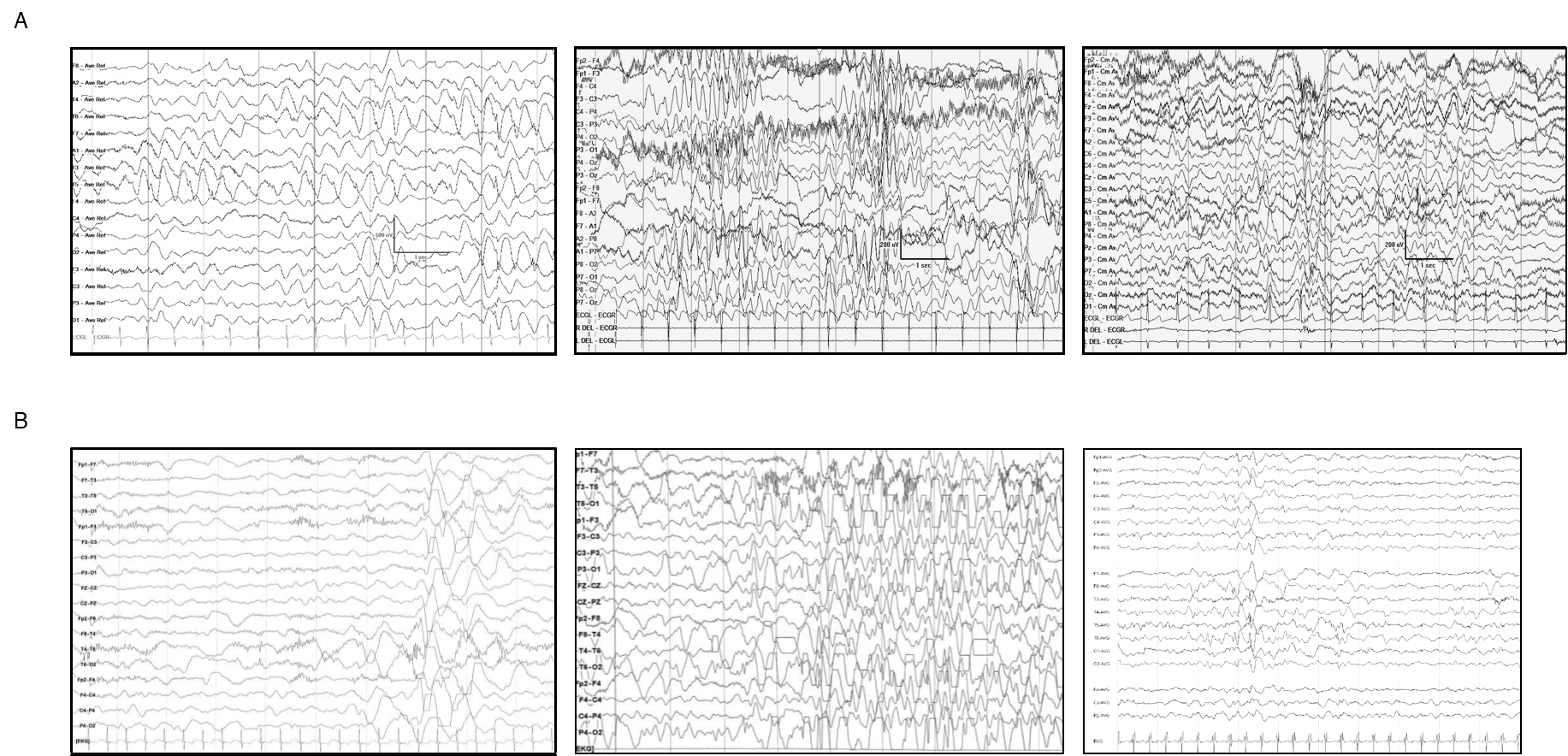

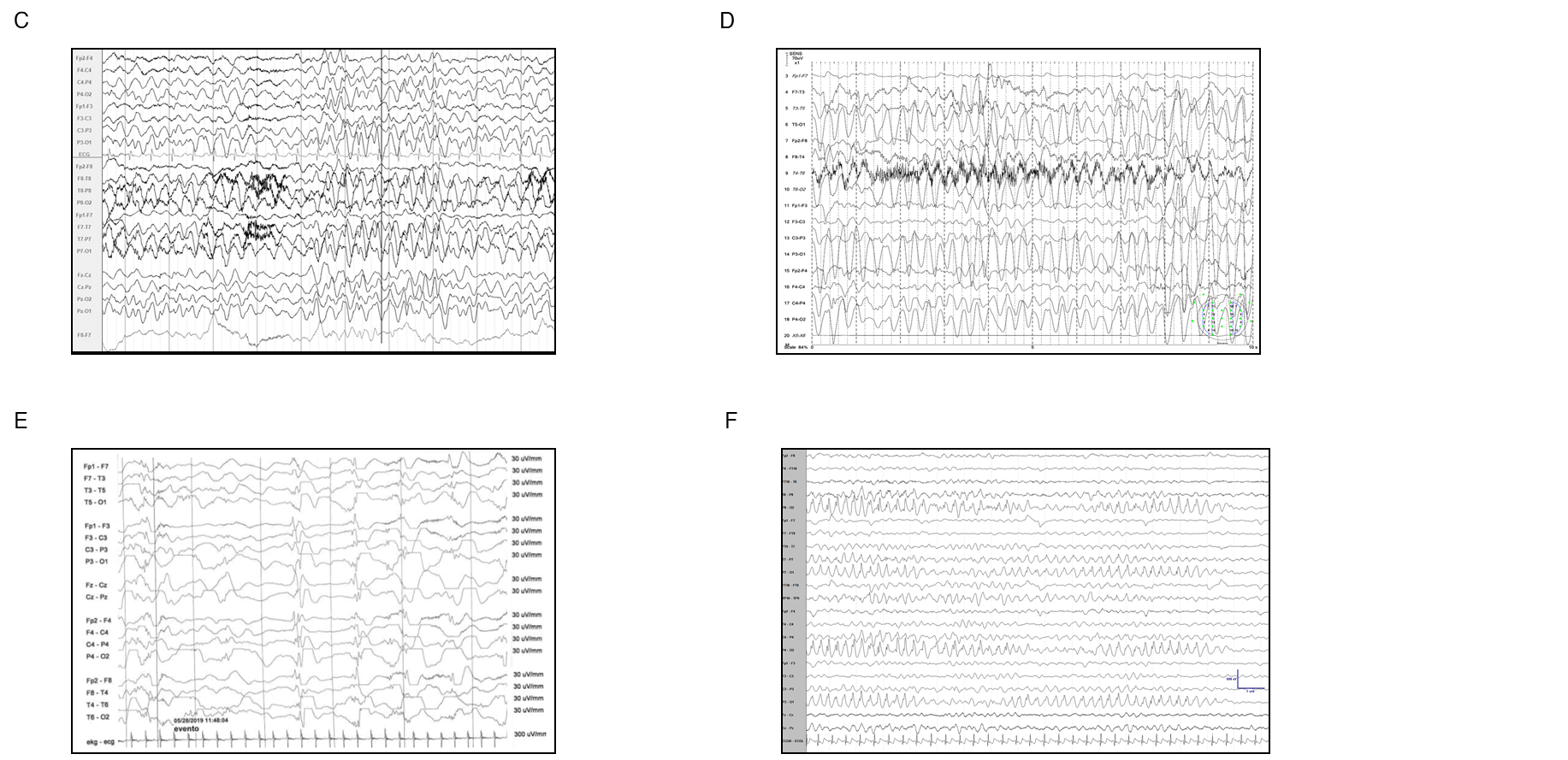

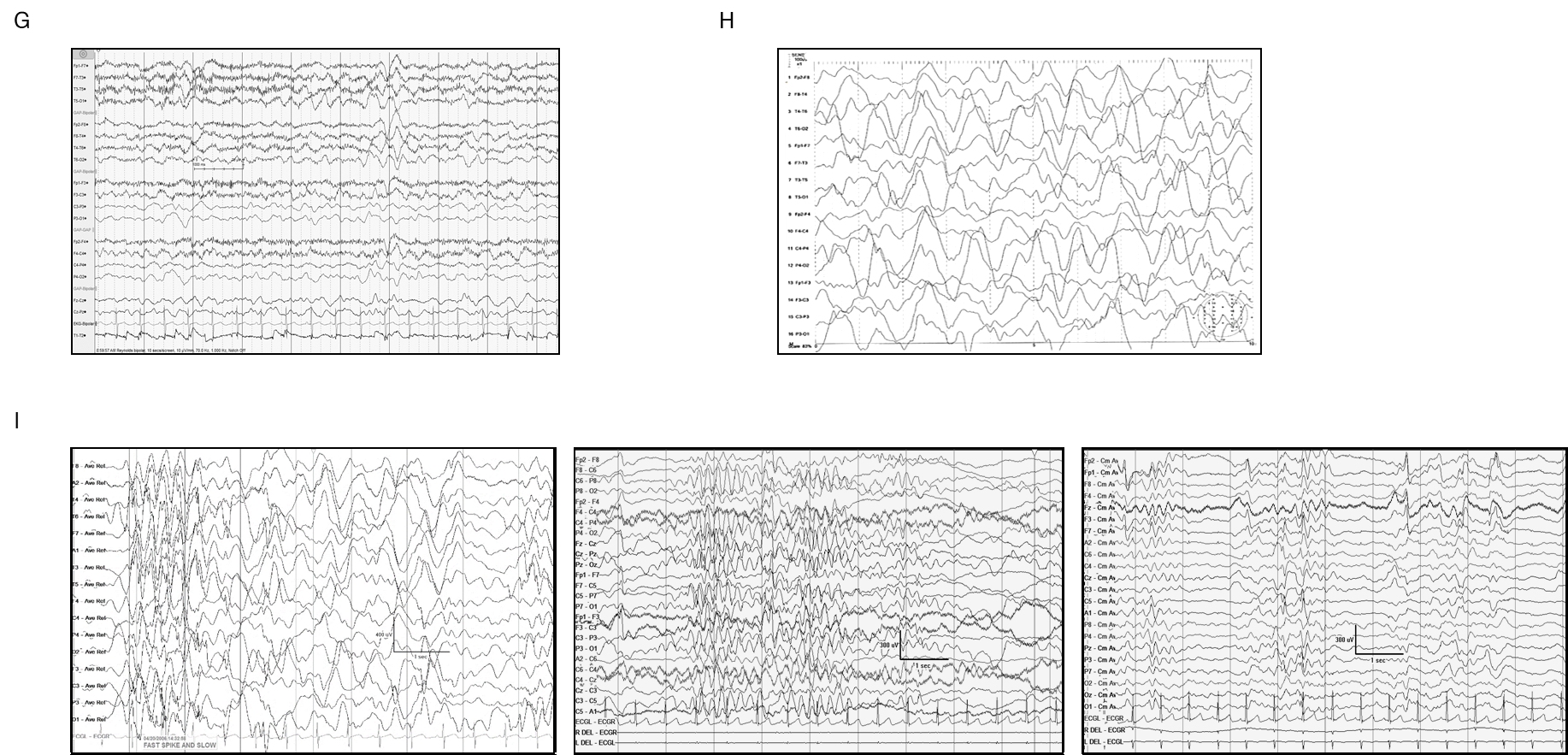
